# Worldwide clustering and infection cycles as universal features of multiscale stochastic processes in the SARS-CoV-2 pandemic

**DOI:** 10.1101/2021.12.20.21268095

**Authors:** Marija Mitrović Dankulov, Bosiljka Tadić, Roderick Melnik

**Affiliations:** Department of Theoretical Physics, Jožef Stefan Institute, Jamova 39, Ljubljana, Slovenia; Complexity Science Hub, Josefstaedter Strasse 39, Vienna, Austria; MS2Discovery Interdisciplinary Research Institute; M2NeT Laboratory and Department of Mathematics, Wilfrid Laurier University, Waterloo, ON, Canada; BCAM - Basque Center for Applied Mathematics, Alameda de Mazarredo 14, E-48009 Bilbao, Spain

## Abstract

Predicting the evolution of the current epidemic depends significantly on understanding the nature of the underlying stochastic processes. To unravel the global features of these processes, we analyse the world data of SARS-CoV-2 infection events, scrutinising two eight-month periods associated with the epidemic’s outbreak and initial immunisation phase. Based on the correlation-network mapping, K-means clustering, and multifractal time series analysis, our results reveal universal patterns, suggesting potential predominant drivers of the pandemic. More precisely, the Laplacian eigenvectors localisation has revealed robust communities of different countries and regions that then cluster according to similar shapes of infection fluctuations. Apart from quantitative measures, the immunisation phase differs significantly from the epidemic outbreak by the countries and regions constituting each cluster. While the similarity grouping possesses some regional components, the appearance of large clusters spanning different geographic locations is persevering. Furthermore, cyclic trends are characteristic of the identified clusters, dominating large temporal fluctuations of infection evolution, which are prominent in the immunisation phase. Meanwhile, persistent fluctuations around the local trend occur in intervals smaller than 14 days. These results provide a basis for further research into the interplay between biological and social factors as the primary cause of infection cycles and a better understanding of the impact of socio-economical and environmental factors at different phases of the pandemic.

## Introduction

In social dynamics, the genesis of a collective phenomenon arising from contagious social interactions involves mechanisms of self-organised criticality^1,2^. It depends on each individual involved, based on its actual contacts, psychology and behaviour. In the presence of viruses, these mechanisms are additionally shaped by firm biological factors. Recent developments of SARS-CoV-2 pandemic^3,4^ revealed a specific global phenomenon emerging from the stochastic multi-scale processes. The infection incidence occurs with a high temporal resolution at the interactions between the virus and human hosts, whose biological features and social behaviours significantly contribute to the epidemic’s spreading^5^. At the molecular scale, the virus-host interactions^6–8^ crucially depend on the virus biology and genetic factors determining the host’s immunity towards the virus in question^9,10^. Thus, the occurrence of an infection event and the infection manifestation may lead to a range of different scenarios from asymptomatically infected to severe health issues and fatalities^11–14^. Multiple other factors may play a role^15^, depending on the population genetic features and social life^16^. They include cultural, political and economic aspects, official and spontaneous reaction to the crisis, and the organisation of the health care system, all of which may significantly differ between different geographical locations^17^. Moreover, the actual impact of these factors changes over time as the epidemic develops, in particular, since the appropriate vaccines targeting SARS-CoV-2 viruses^1,18^ are available, thus enabling massive immunisation of the population. Attempts were made to identify different parameters that may influence the epidemic and estimate their mutual interdependence and impact. For example, the human-development index, built-up-area-per-capita, and the immunisation coverage appear among the statistically high-ranking drivers of SARS-CoV-2 epidemic^15^.

In addition, temporal variations occur at all scales, from the virus mutations^8^ to changed behaviours of each individual and population groups, e.g., due to the government imposed measures^4,19^, or adaptation caused by the awareness of the current epidemiological situation^20,21^. These variations increase the stochasticity of the infection and contact processes, making the prediction of their output even more difficult. For real-time epidemic management and the predictions of further developments, it is crucial to understand the nature of the underlying stochastic processes and the factors that can significantly influence them. For this purpose, the empirical data analysis and theoretical modelling^22^ provide complementary views of these complex processes. For example, agent-based models capture the interplay of the bio-social factors at the elementary scale of the virus-host interactions at high temporal resolution^5,23–31^. On the other hand, more traditional compartmental models^32^ consider a coarse-grained picture of the population groups having different roles in the process. Another research line aims at the mathematical description of the exact empirical data, in particular, for the outbreak phase^33,34^. For instance, different studies provided tangible arguments for the cause of the changing shape of the infection curve comprising the appearance of linear and power-law segments^35,36^, prolonged stagnation periods, and multiple waves^37^. Since the beginning of the epidemic, empirical data were collected over different countries or provinces^38^. Despite the coarse-grained spatial and temporal structure (daily resolution), these data may contain relevant information about the temporal aspects of the epidemic at different geographical locations. Previous studies, based on the empirical data regarding the dynamics of interacting units in many complex systems, provided valuable information about the related stochastic processes. Some striking examples across different spatial and temporal scales include the influence of the world financial index dynamics on different countries^39,40^, traffic jamming^41,42^, brain-to-brain coordination dynamics^43,44^, and the cooperative gene expressions along different phases of the cell cycle^45,46^. Similarly, the collected data of SARS-CoV-2 spreading enable a possibility to investigate the infection dynamics in various details and more appropriate modelling of the emergent behaviours. In this respect, a larger-scale picture may emerge by studying temporal fluctuations of the world infection dynamics. More subtle questions regard the indicators for hidden mechanisms arising from the interplay of the above-mentioned biological factors and different social behaviours^5,21,23,47,48^ behind the observed epidemic development.

In this work, we focus on some of these critical issues by studying the data that are publicly available at GitHub^38^ collected over different countries or regions (provinces). Using the datasets of the daily recorded number of confirmed infection cases, we consider two separate segments of time series. Namely, the records for the first eight months of the epidemic, starting from the first registered case in each country, represents the epidemic’s *outbreak phase*. Meanwhile, the last eight months (preceding the data collection on September 30, 2021), during which the pharmaceutical intervention was available in most of the countries, characterises the initial *immunisation phase* of this pandemic. Our quantitative analysis comprises three levels of information: the network mapping and spectral analysis, K-means clustering of pairs of time series, and detrended fractal analysis of individual time series. In addition to quantifying the differences between the outbreak and immunisation phase, our results reveal two global features of the SARS-CoV-2 pandemic. Firstly, the worldwide groups of countries (and provinces) robustly appear in clusters having a similar temporal evolution of the infection dynamics. This clustering suggests that the environmental and socio-economical factors and government-imposed measures can certainly influence small-scale fluctuation characteristics of the clusters but do not significantly change the course of the process on larger scales. Secondly, the epidemic evolution exhibits ubiquitous waves driven by the cyclic infection dynamics, where several typical cycles appear associated with the identified clusters. Again, the shape of these typical cycles coincides with the mentioned clustering mechanisms. Hence, their origin and potential control will remain challenging within purely social measures. A more detailed analysis of the complex feedback between biological and social factors at all scales is needed.

## Results

As stated in the Introduction, we consider worldwide recorded time series of the infection cases. Defining two distinct eight-month periods in the epidemic’s evolution is motivated by the appearance of SARS-CoV-2 vaccines in the latter period, enabling pharmaceutical intervention measures not available in the outbreak phase. For illustration, a few examples of time series recorded in different countries are shown in Fig. 1.

**FIG. 1:**
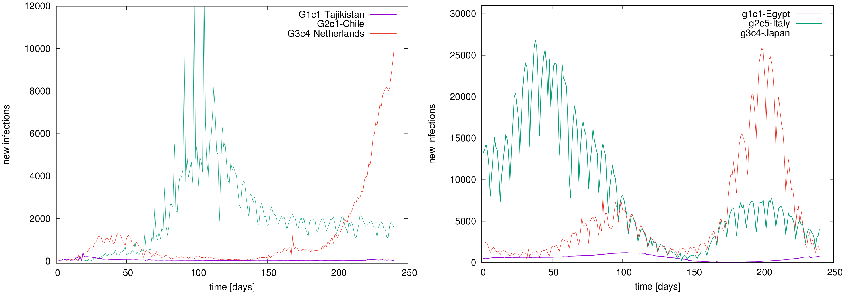
Examples of time series. Temporal evolution of confirmed infection cases in different countries, belonging to different groups in the outbreak (left) and immunisation phase (right).

### The correlation networks mapping in the outbreak and immunisation phase

The network mapping is based on the cross-correlation coefficient *C*_*ij*_ of the pairs of time series{ *i, j* } and a suitably selected threshold. Hence, the correlations exceeding the threshold *θ* are accepted, making the adjacency- matrix elements *A*_*ij*_(*θ*) = Θ(*C*_*ij*_ − *θ*) − *δ* _*ij*_ of an undirected unweighted network. Before selecting the threshold, a filtering procedure was applied to the complete correlation matrix to enhance the positive correlations of interest in this work (see Methods). The applied methodology was proved useful in quantifying correlations of time series in diverse type of data^39,41–46^. Figure 2a shows probability distributions of filtered correlations coefficients for the outbreak and immunisation period. While both probability distributions have a peak at a value *C*_*ij*_ *<* 0, they have slightly different shapes. They both have a pronounced tail for positive values of correlation coefficients, where the distribution *P* (*C*_*ij*_) for the outbreak period has a slower decay at correlations *C*_*ij*_ *>* 0.2. The appropriate threshold is selected considering changes in spectral properties of the adjacency matrix with the increasing threshold, as explained in the following. Figure 2b shows the Kolmogorov-Smirnov (KS) distance between the eigenvalues of the *A*_*ij*_(*θ*) compared to the one at *θ* = 0 depending on the threshold *θ* for the outbreak and immunisation networks. We see that the KS distance grows slowly with *θ* up to the value ∼0.4; meanwhile, the growth becomes rapid for the values of *θ >* 0.5 for both networks, suggesting a profound change in the networks’ structure when the threshold exceeds *θ* = 0.5. Thus, we select this turning point as the optimal threshold value. Moreover, the networks obtained by applying the threshold weight *θ* = 0.5 are sufficiently sparse; meanwhile, their spectral properties do not differ drastically from the corresponding outbreak and immunisation period networks at *θ* = 0 containing all positive correlations. The resulting networks for *θ* = 0.5 are visualised in Fig. 3. See also Supplementary Information (SI) for more details.

**FIG. 2:**
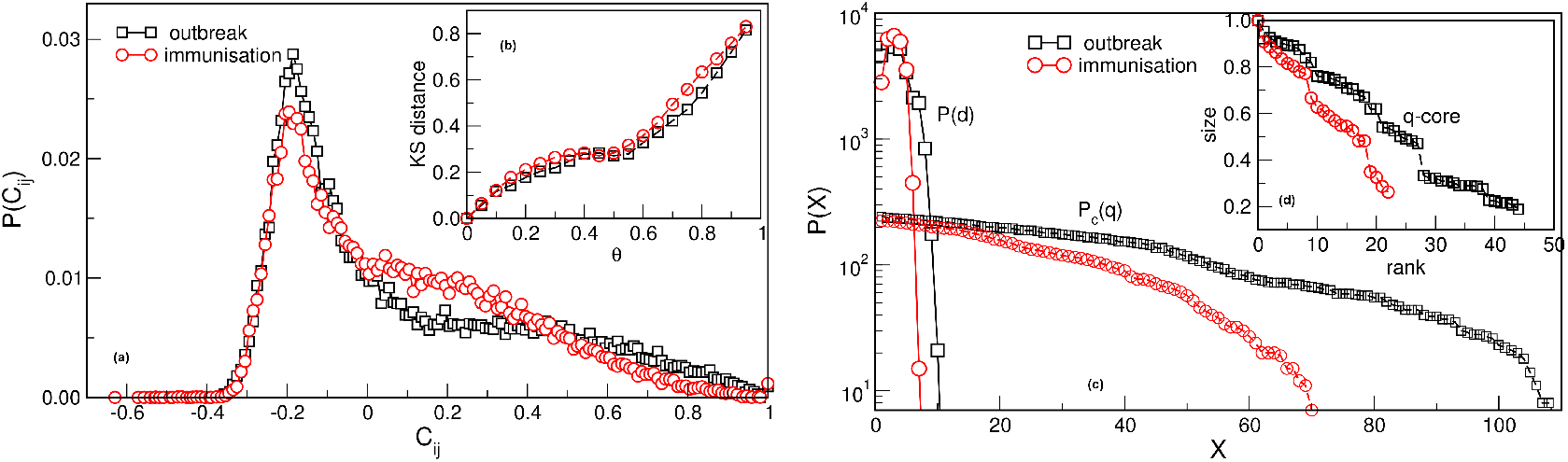
Network measures. (a) The normalised probability density function of the filtered correlation coefficients for the outbreak and immunisation periods. (b) The Kolmogorov-Smirnov distance between the eigenvalue spectrums of networks obtained for *θ* = 0 and different values of *θ* > 0, plotted against *θ* > 0. (c) The distribution of the shortest-path distances P(d) vs the distance d and the cumulative distribution *Pc*(*q)* of the node’s degree *q* for the outbreak and immunisation networks with the threshold *θ* = 0.5. (d) The size of the q-core of these networks plotted against the q-rank.

**FIG. 3:**
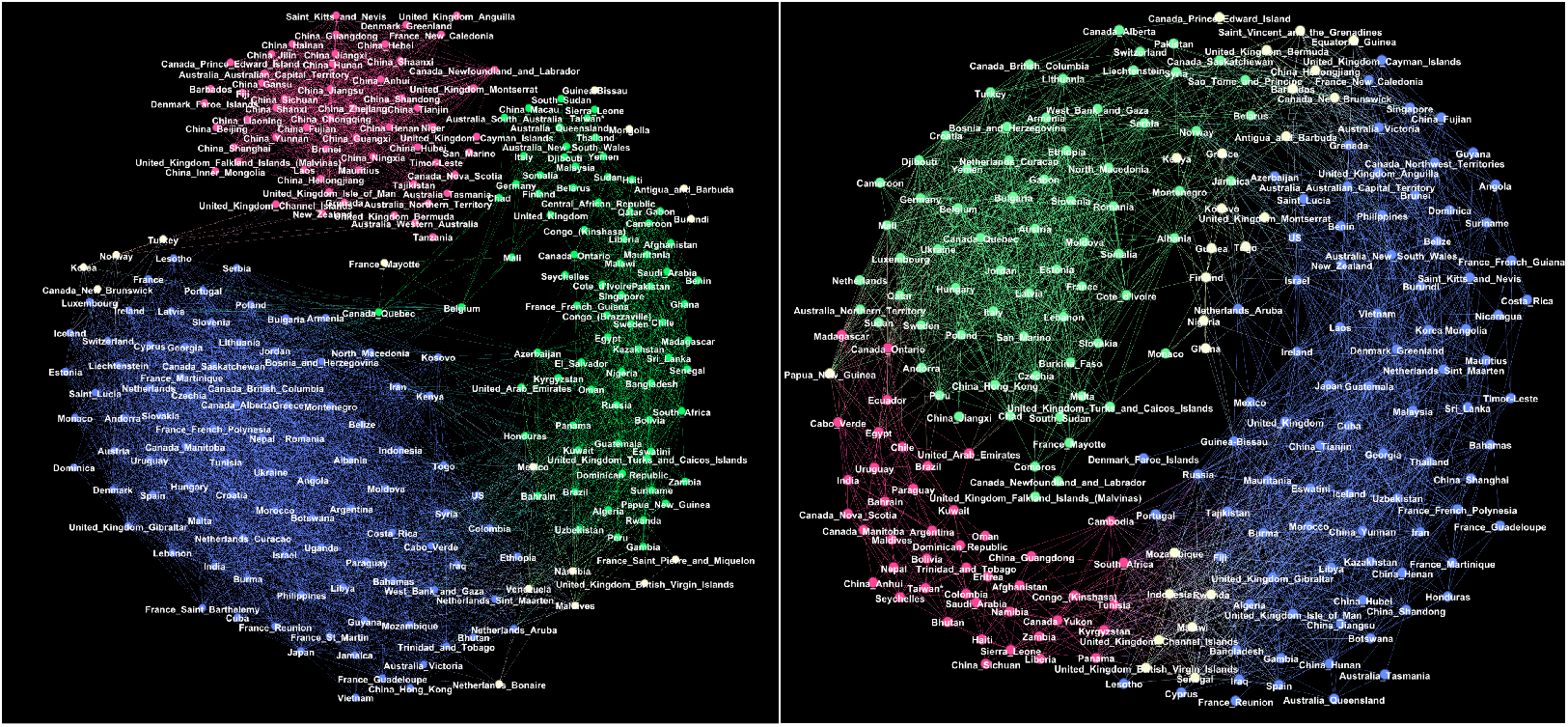
Giant connected components of the correlation networks at the threshold *θ* =0.5 for the outbreak period (left) and the immunisation period (right). Red, green and blue colours indicate groups of nodes in three respective communities G1, G2, G3 in the outbreak network, and g1, g2, g3 in the immunisation period network, determined by the eigenvector-localisation, see text and Fig. 4. Unclassified borderline nodes are shown in white colour. Labels on nodes identify the corresponding country or province. The complete lists of nodes in each community are given in Tables S1-S6 in Supplementary Information.

The giant connected component of each network exhibits a community structure, i.e., the occurrence of groups of nodes that are better connected among themselves than with the nodes outside that group, cf. Fig. 3. The identity of nodes comprising each community is determined using the localisation of the eigenvectors associated with the three lowest nonzero eigenvalues of the normalised Laplacian operator^49^, as explained in Methods^50^. The eigenvalues of the normalised Laplacians for two networks are shown in ranking order in Fig. 4, middle panel. Several lowest nonzero eigenvalues appear to be separated from the bulk in both networks. This network feature is compatible with the existence of mesoscale communities, on which the corresponding eigenvectors tend to localise^49,50^. The scatterplots of the eigenvectors associated with three lowest nonzero eigenvalues, see Fig. 4, show three differentiable branches, here indicated as *G*1, *G*2, *G*3 for the outbreak, and *g*1, *g*2, *g*3 for the immunisation phase network. The indexes with a nonzero component of the eigenvectors in each branch mark the IDs of the nodes belonging to the corresponding community. The complete lists of nodes in each community (group) are given in Tables S1–S6 in SI.

**FIG. 4:**
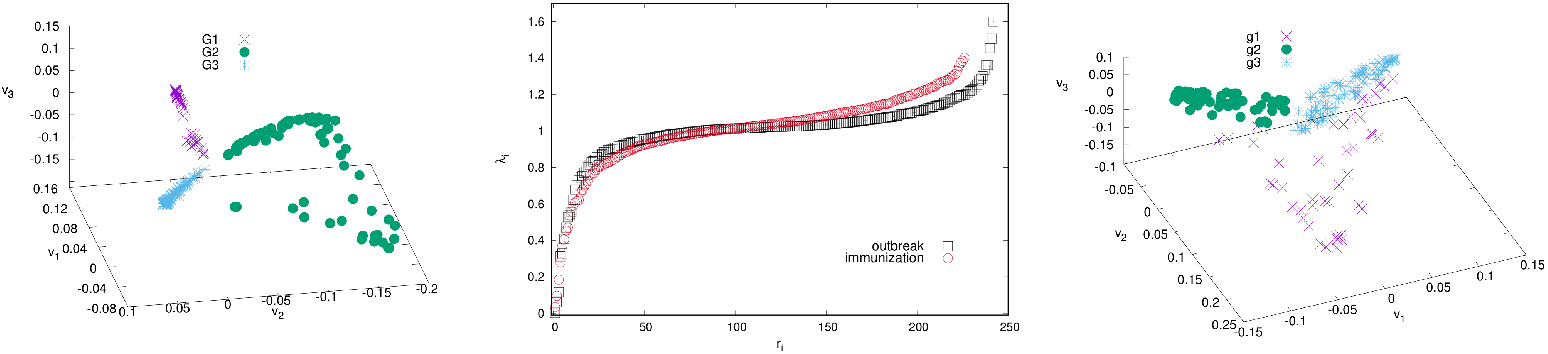
The ranking of eigenvalues of the normalised Laplacian for the outbreak and immunisation networks (middle). Scatter plots of the eigenvectors *v*1,*v*2,*v*3 corresponding to the three smallest non-zero eigenvalues for the outbreak (left) and immunisation period networks (right). Branches indicated by different colours identify the communities (groups) of nodes of the corresponding network in Fig. 3.

Even though both networks exhibit three major communities, the structural differences between the two networks in Fig. 3 are apparent. They indicate the corresponding differences in the fluctuations of the infection rates in the world regions during the immunisation phase, compared to the epidemic’s outbreak, when the whole population was practically susceptible to the infection. These differences are quantified by several graph measures, see Fig. 2a-d and Table I. Compatible with these graph-theory measures are the span of the exponentially-decaying degree distributions *P*_*c*_(*q*) and different distributions of the shortest-path distances *P* (*d*), shown in Fig. 2c. We also show the prominent differences in the q-core structure of these networks, cf. Fig. 2d. More importantly, the majority of nodes that belong to the same community in the outbreak phase network appear to be a part of entirely different communities in the immunisation phase network, cf. Fig. 3 and the corresponding lists in Supplementary Information. More precisely, we find that only 625 edges established in the outbreak phase persist in the immunisation phase network. They are shown in Fig. S2 left, in Supplementary Information. A more systematic comparison is made by computing the overlap (Jaccard index defined in Methods) for the correlation networks determined from the successive two-month intervals, see Fig. S2, right. The overlap systematically remains below 15%, suggesting that the fluctuation patterns at these intervals can vary between the countries or even provinces within the same country.

**Table I:** For the outbreak and immunisation period networks: the number of nodes *N*, edges *E*, and triangles the graph diameter D and density ρ; the average path length < ℓ >, degree < *q* >, and clustering coefficient < *C*_*c*_ >.

| | $N$ | $E$ | $D$ | $\langle \ell \rangle$ | $\langle q \rangle$ | $\langle C_c \rangle$ | $\#\Delta$ | $\rho$ |
| --- | --- | --- | --- | --- | --- | --- | --- | --- |
| outbreak | 243 | 9284 | 11 | 3.55 | 38.2 | 0.735 | 58508 | 0.158 |
| immunis. | 227 | 5660 | 7 | 3.10 | 24.9 | 0.656 | 18857 | 0.112 |

### K-means clustering and multi-fractality of time series within identified communities

To further explore the nature of temporal fluctuations of the infection time series of the countries and provinces within each community found using spectral analysis, we apply the K-means algorithm adapted for time series analysis^52^, see Methods. It appears that each topological community is further partitioned into several clusters, for example, *G*1*c*1 … *G*1*c*4, for the group *G*1, and so on. Inside each cluster, the corresponding time series have a similar evolution pattern. Hence, the cluster’s typical time series (centroid) is determined for each identified cluster. The results are shown in Fig. 5 both for the outbreak and immunisation phase; in the figure legends, the number of countries or provinces belonging to a given cluster is indicated in the brackets in each panel. The names of countries and provinces belonging to each cluster in each group are given in Tables S1 to S6 in Supplementary Information. Notably, in each network’s group, there is one large and one medium-size cluster. Meanwhile, there are several single-country centroids; as a rule, they indicate a large-population country.

**FIG. 5:**
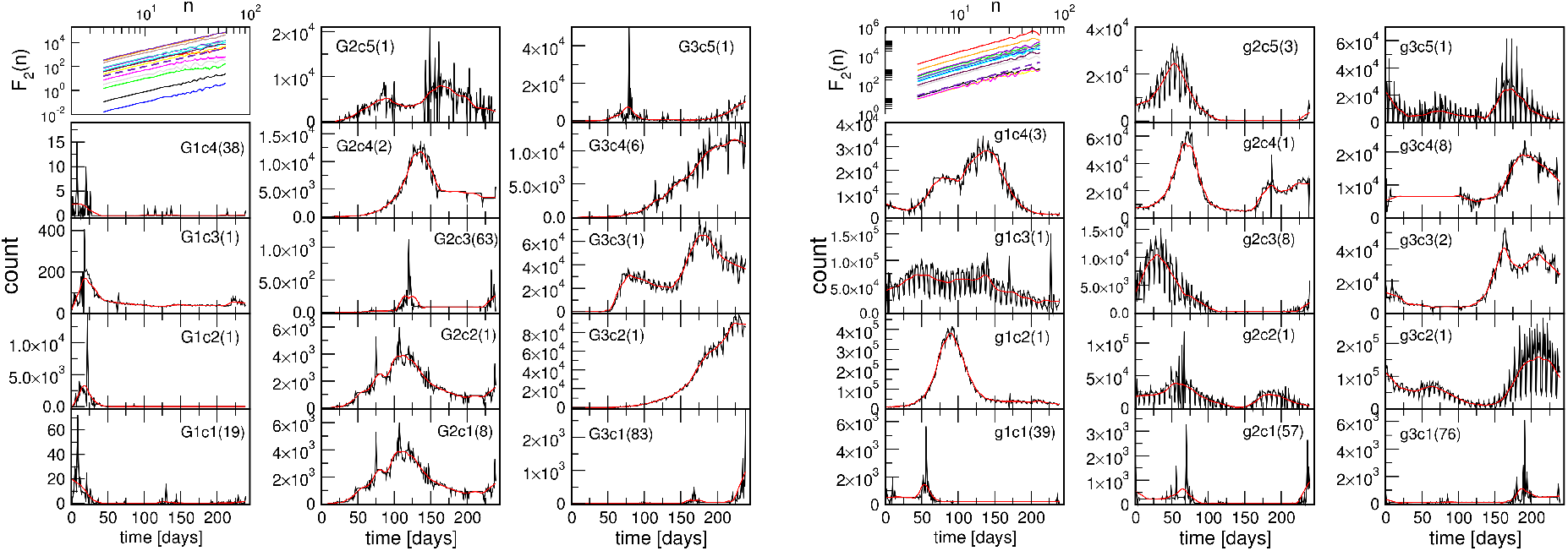
Centroids of clusters c1 to c5, found for three groups G1, G2 and G3, in the outbreak phase network (left three columns), and groups g1, g2 and g3 in the immunisation phase network (right three columns). In each panel, the number of countries and provinces belonging to that cluster is shown in brackets; the smooth red line represents the centroid’s trend. The top left panel in each figure shows the fluctuations function F2(*n*) vs segment length n for the identified trends; the slope *h*2 = 2 is indicated by the dashed line.

Next we consider the fluctuation function *F*_2_(*n*) vs the interval length *n* for each time series separately, see some examples in Fig. 6, and Fig. S4 in SI. We realised that the similarity of the time series belonging to each cluster manifests itself in the apparent similarity of the slopes of their fluctuation function, which defines the corresponding Hurst exponent. As Fig. 6 shows, two different slopes of the fluctuation function can be identified for a majority of time series. At the intervals *n <* 14, a Hurst exponent 0.5 ≲*h*_2_ ≲1 can be determined, indicating persistent fluctuations occurring at these time intervals. Meanwhile, an exponent *h*_2_ *>* 1, characteristic to the fractional Brownian motion, is found for *n* ≥14. In some cases, the determined Hurst exponent reaches values close to two. The histograms of the observed Hurst exponents are shown in Fig. 6d,e. Compatible with the grouping and different shapes of centroids in the immunisation phase, the distributions of lower and higher values of the Hurts exponents are also different with the increased incidence of the value *h*_2_ ≂ 0.5 (white noise), and *h*_2_ ≂ 2 (periodic signals) in the immunisation phase. In the following, we show that these large values of the Hurst exponent in many of the studied time series can be related to the occurrence of *cyclic trends*.

**FIG. 6:**
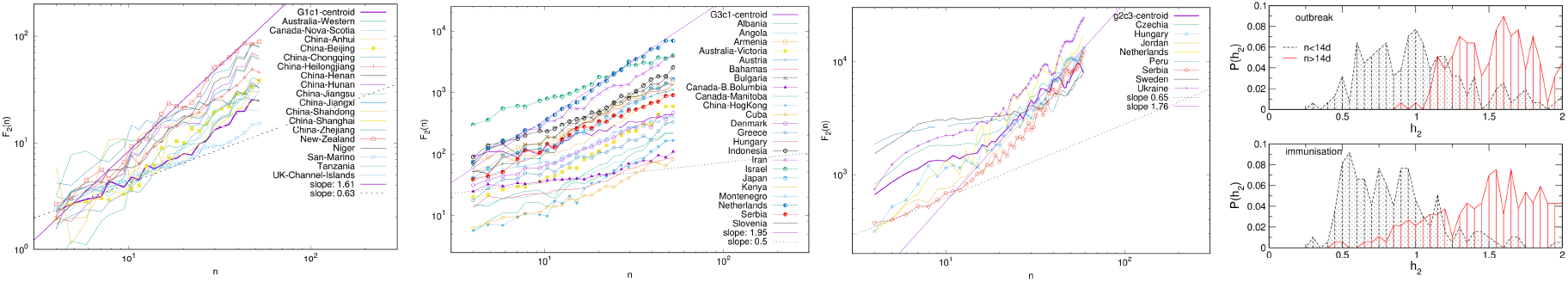
Examples of the standard deviation F2(*n*) of time series vs the interval length *n* for K-means clusters G1c1 and G3c1 identified within topological communities G1 and G3 in the outbreak (a,c), and cluster g2c3 in the immunisation phase networks (c). The distribution of the measured Hurst exponents for the intervals n < 14 days and n ≥ 14 days in the outbreak and immunisation phase (d,e).

Two prominent examples are shown in Fig. 7. The methodology of determining local trends in these time series is described in Methods. The original time series shows a cyclic trend, where the cycle length can vary from region to region. A separate analysis of the fluctuation functions for the trend and the fluctuations around the local trend (detrended signal) reveals that the trend drives the fluctuations beyond the intervals of approximately 14 days; see the insets to Fig. 7. The trend has true cyclic fluctuations (the Hurst exponent equals two, within error bars) in the range up to *n ≲* 30 days. Meanwhile, beyond this range, both the original signal and trend have a lower Hurst exponent in the range *h*_2 ≳_ 1, characterising a fractional Brownian motion. By extending a similar analysis to the above-mentioned typical time series (centroids), we find that they also exhibit cyclic trends but with different cycles characterising different clusters of countries. The corresponding trends are also shown in each panel of Fig. 5 as a red line on the top of the related centroid. The trend fluctuation functions *F*_2_(*n*) vs *n* shows the cycle characteristics in a large range of the intervals *n*, cf. top left panels of Fig. 5. They differ from cluster to cluster and, even for the same country, the cycles also differ in the outbreak and immunisation phase. Generally, larger cycles (in the length and amplitude) are observed in the immunisation phase as compared to the outbreak period, cf. Fig. S5 in SI. Remarkably, these findings imply that the cycles (or the infection waves) represent an inherent feature of current pandemic which may have some long-lasting consequences.

**FIG. 7:**
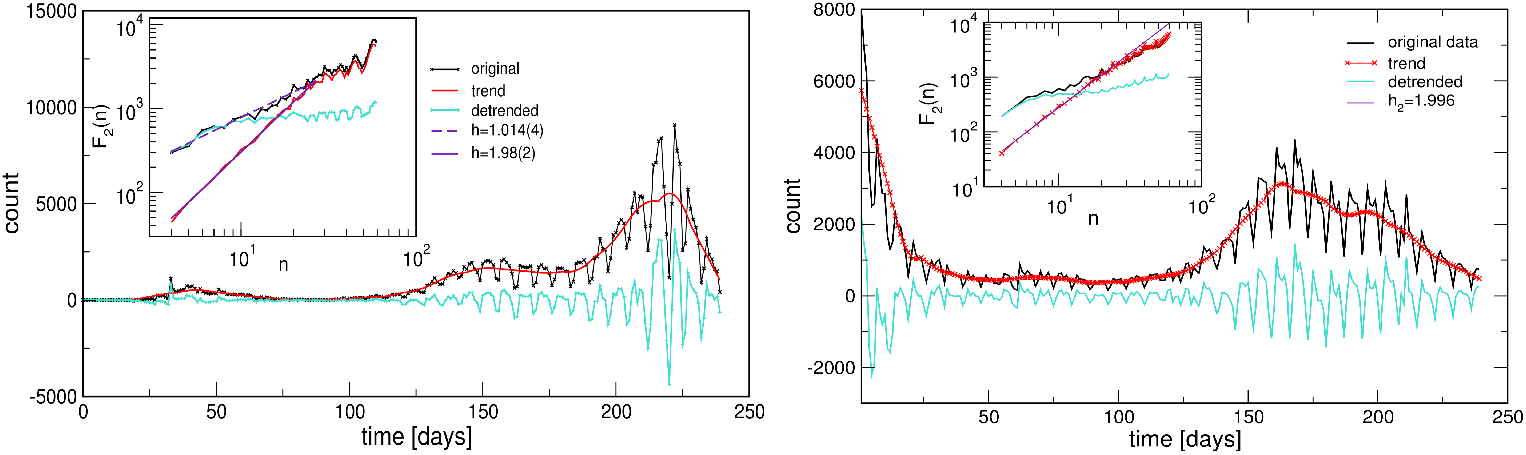
Two examples of the infection time series showing cyclic trends during the outbreak phase (from Israel, left), and during the immunisation phase (from Portugal, right). Insets show the corresponding functions of the standard deviation fluctuations for the identified trends and the original and detrended time series.

## Discussion and Conclusions

We have analysed the worldwide empirical data of SARS-CoV-2 epidemic^38^, focusing on the new infection dynamics with a daily resolution. As explained above, the data are purposefully divided into two periods, corresponding to the epidemic’s outbreak and the initial immunisation phase, respectively. Three complementary methods of quantitative analysis have been performed. Specifically, we have analysed the mesoscopic structure of the networks, which embody the significant pairwise correlations among the infection time series of different countries or provinces. The further similarity in the pairs of time series has been analysed by K-means clustering. Finally, the fluctuation function of each time series has been determined using the detrended time series analysis. Our analysis has revealed global clustering and several universal features of the infection dynamics. Our main conclusions are summarised here:

- *the worldwide clustering* reveals significant similarity in the temporal patterns of infection that exceeds strictly geographical regions;
- *the cyclic trends* dominate the infection fluctuations, implying the prevalent infection waves and multi-scale fluctuations around these cycles; some typical cycles were determined in conjunction with the identified clusters;
- *the immunisation phase differs* in all measures from the epidemic outbreak phase, thus quantifying the impact of the (currently partial) immunisation coverage onto the course of the pandemic.

The mesoscopic (community) structure, as shown in Fig. 3 is one of the striking characteristics of the infection-correlation networks; remarkably, it occurs already at zero thresholds, see Fig. S1 in SI. What comes as a surprise is that these communities constitute almost entirely different nodes (countries or provinces) in the immunisation phase compared to the outbreak phase. Only a few edges established during the outbreak phase persist throughout the entire evolution of the epidemic, as shown in Fig. S2 in SI. Consequently, the same applies to the contents of the clusters found in these two phases, cf. Tables S1-S6 in SI. Notably, a given geographic location and potentially similar cultural and economic development levels, similar healthcare systems and other related factors play some role. However, even such regional groups appear to be a part of a worldwide cluster in both representative phases of the pandemic. Such a picture probably emerges under another dominant driver, common to countries at different locations, and with different cultural and economic developments. In this context, the biology factors, the virus mutations in the interplay with the social behaviour of individuals and groups in the crisis seems to be of the primary importance for the genesis of sustained infection waves, quantified by cyclic trends in different clusters, cf. Fig. 7. Our analysis suggests that the waves are ubiquitous in all countries and regions in both representative phases of the pandemic. Meanwhile, the timing, duration and amplitude of these waves vary between different clusters of countries and provinces, likely depending on the applied measures and the corresponding variations in the population behaviours. Moreover, the small-scale fluctuations around these cyclic trends seem to be more region-specific, and depending on the immunisation measures; two comparative examples are shown in Fig. S5 in SI. A more systematic analysis of these fluctuations and the impact of the immunisation level on the infection dynamics merits future study.

Our analysis of the world infection dynamics of the SARS-CoV-2 pandemic revealed several universal features of the underlying multiscale stochastic processes that go beyond the geographical impact, locally-imposed governmental measures, and partial immunisation phases. Indeed, while these measures are truly valuable for short-term effects, saving lives, and maintaining the functional healthcare system in each country^19^, they are much less effective in changing the fundamental nature of the infection process, rooted in the interplay of biology and social behaviours. This work has provided an in-depth analysis of the pandemic’s fundamental phases with an overview that can guide further research into the nature of biosocial interdependencies. The latter factor plays a critical role in the SARS-CoV-2 evolution, where individual biological features of the participants and their role in the collective behaviours need to be better understood. Our effective long-term management of the pandemic and prediction of its future developments rely upon our ability to continue unfolding critical attributes of the underlying biosocial stochastic dynamics.

## Data & Methods

### Data acquisition, preparation, and mapping

We consider the worldwide data of the number of new infection cases downloaded from GitHub^38^. The dataset contains the number of daily detected new cases for 279 countries or provinces. For this work, we select time series in two eight-mount periods comprising the epidemic’s outbreak phase (starting from the first registered case in a given country or province) and the immunisation phase (22 January 2020 until 30 September 2021). The corresponding number of countries and provinces with the active epidemic’s data traced in both periods is 255. For instance, the first case in France was detected on 24 January 2020, and thus the outbreak time series covers the period from that date until 19 September 2020. However, Slovenia had the first registered case on 5 March 2020; hence its outbreak time series cover 5 March until 30 October 2020. Meanwhile, the immunisation period is from 3 February 2021 to 30 September 2021, equal for all considered countries and provinces.

By mapping these datasets, we obtain two correlation networks for the outbreak and immunisation phase, respectively, where the network’s links stand for significant positive correlations. We first compute the Pearson’s correlation coefficient for the corresponding pairs (*i, j*) of the time series

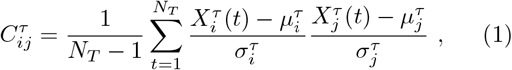

where *τ ∈*{*O, V* }, 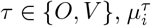 is average value of the time series of country *i* during period *τ*, and 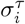 is standard deviation of time series 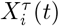. To remove spurious correlations, we apply the filtering procedure standardly used in these type of network mapping^41,43,45^. More precisely, the matrix elements 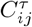 are first transformed to the interval [0, 1] by 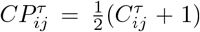, and then multiplied by a factor 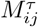 which is obtained in the following way. From the rows *i* and *j*, the diagonal elements are removed and the considered elements 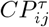 and 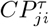 are placed at the beginning of the row *i* and *j*, respectively, thus obtaining two *n* = *N* − 1 dimensional vectors 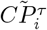 and 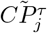. Then 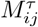 is computed as Pearson’s coefficient between these two vectors. The matrix element of the filtered correlation matrix 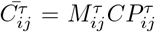 is then mapped back to the interval [− 1, 1]. Finally, the elements of the network’s adjacency matrix are defined as 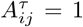 when the matrix elements 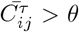 exceed a specified threshold value *θ*, and zero otherwise. The threshold value *θ* is determined concerning the network’s spectral properties, as described below.

### Network’s spectral analysis and community detection

The above-described data mapping should lead to undirected unweighted networks; the nodes represent countries (or provinces), and links indicate the positive correlations between infection incidences exceeding a threshold *θ*. We use the spectral properties of networks to obtain the adequate threshold value, where the guiding criteriums are the network’s sparseness and the relative stability of the community structure. Starting from *θ* = 0, we increase it by the value 0.05 and solve the eigenvalue problem of the corresponding adjacency matrix, **A***v*_*i*_ = *λ*_*i*_*v*_*i*_| _*θ*_, and calculate the spectrum {*λ*_1_, …, *λ*_*N*_ }_*θ*_ for each threshold *θ*. Comparing the network spectrum for *θ* = 0 with one obtained for the network for each *θ >* 0, we determine the Kolmogorov-Smirnov distance and plot it against *θ*; we determine a characteristic threshold *θ* common to both networks, as shown in the text.

We study the community structure of the networks for the outbreak and immunisation period using spectral analysis and the eigenvalue problem of the normalised Laplacian related to the network’s adjacency matrix. In mathematics theory^50**?**^, the number of smallest non-zero eigenvalues of the Laplacian matrix is a good indicator of the number of communities. The matrix elements of the normalised Laplacian for undirected binary network represented by the adjacency matrix **A** are defined as

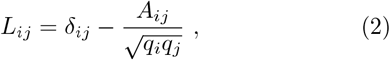

where *q*_*i*_ and *q*_*j*_ are degrees of nodes *i* and *j*. For the normalised Laplacian (2), we solve the eigenvalue equation 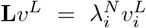 and determine all eigenvalues and eigenvectors. In the case of a connected network, these eigenvalues are non-negative. One zero-eigenvalue appears with strictly positive eigenvector’s components^49^. Consequently, the orthogonal eigenvectors corresponding to the three smallest non-zero eigenvalues localise on the communities of the network. Hence, the scatter plot of the components of these eigenvectors shows a branching structure. Each branch contains indexes of the non-zero eigenvector components, that is, the nodes belonging to a network’s community^50^.

The size of the q-core of the networks is determined by removing the nodes with the increasing degree *q*. Several other graph properties are determined, and the networks are visualised using *Gephi* software^51^.

### K-means clustering of time series

The implementation of the K-means algorithm for clustering of time series in Python known as *tslearn*^52^ is used. K-means is an unsupervised machine learning algorithm that aggregates data points according to similarities, starting with *K* randomly positioned centroids. Based on these centroids, data points are assigned to the centroid closest to that data point according to some distance metric. The algorithm consists of a certain number of iterative (repetitive) calculations used to optimise the positions of the centroids. Considering each time series of length *N*_*T*_ as a data point in *N*_*T*_ dimensional space, the appropriate measures enable calculating the distances between these data points. We use the Dynamic Time Wrapping (DTW) algorithm to align time series with centroids and measure their similarities. The DTW is widely used for time series comparison and classification. It performs an optimal alignment between two time series by matching the indices from the first time series to the second time series, subject to several constraints. Specifically, the mapping of indices from the first series to the second series must be monotonically increasing. For the indices *i > j* from the first time series, there must be two indices from the second series *l > k* such that *i* is matched with *l* and *j* is matched with *k*. Meanwhile, the first index from the first series must match the first index of the second time series, and similarly, the last index from the first series must be matched to the last index of the second time series, but these points may have more other matches. The optimal alignment is the one that satisfies all of these restrictions with the minimal cost, where cost is the sum of absolute differences of values for each matched pair of indices. The DTW distance in the K-means algorithm is the value of cost. We use the K-means algorithm with DTW distance to cluster time series and find centroids. Each centroid is again a time series that describes the average behaviour of the time series belonging to one cluster.

### Trends and fractal analysis of time series

Temporal fluctuations are studied by the fractal detrended analysis of each time series. For each time series *x*(*k*), *k* = 1, 2, … *T*, the profile 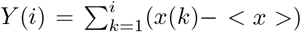 of the time series is divided in *N*_*s*_ segments of the length *n*. The fluctuation function *F*_*q*_(*n*) with the varied segment length *n* is defined as

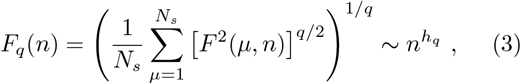

Here, 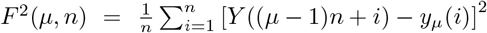 is the standard deviation from a local trend *y*_*µ*_(*i*) on the segment *µ*. For *q* = 2, we determine the Hurst exponent *h*_2_ from the straight-line segments of the log-log plot of the fluctuation function *F*_2_(*n*). For the multifractal analysis, the values of *q ∈* [−4, 4] are varied.

To determine cyclic trends, we use the local adaptive detrending algorithm, see^53,54^, where time series is divided into segments of the length 2*m*+1 overlapping over *m*+1 points. The polynomial interpolation is applied in each segment, and its contribution in the overlapped region is weighted such that it decreases linearly with the distance from the segment’s centre.

## Data Availability

All data produced in the present work are contained in the manuscript

https://github.com/CSSEGISandData/COVID-19/{github}

## Acknowledgments

B.T. work supported by the Slovenian Research Agency (research code funding number P1-0044). M.M.D. acknowledge funding provided by the Institute of Physics Belgrade, through the grant by the Ministry of Education, Science, and Technological Development of the Republic of Serbia. R.M. is grateful to the NSERC and the CRC Program for their support and he is also acknowledging the support of the BERC 2018-2021 program and Spanish Ministry of Science, Innovation, and Universities through the Agencia Estatal de Investigacion (AEI) BCAM Severo Ochoa excellence accreditation SEV-2017-0718, and the Basque Government fund “AI in BCAM EXP. 2019/00432”

## Author contributions statement

B.T., R.M., M.M.D. designed research, M.M.D. collected data, M.M.D., B.T. contributed program tools and performed analysis, B.T., M.M.D., R.M. analysed data, B.T. produced figures, B.T., R.M. wrote the manuscript, all authors reviewed the manuscript.

## Additional information

**Supplementary Information** file uploaded;

## Supplementary Information for

## I. TABLES

Groups of countries and provinces as communities identified by the spectral analysis corresponding to three leading eigenvalues in the connected component of the correlation network for the threshold *θ* = 0.5 for the first eight months (outbreak) and the last eight months (immunisation) networks; these networks are shown in Fig. 3 in the main text:

### A. Outbreak phase Groups G1,G2,G3: clusters

Names of the countries and provinces in the group G1 belonging to four different clusters are shown in Table S1. Similarly, the group G2 consists of five clusters, listed in Table S2, and the five clusters belonging to the group G3 are shown in Table S3.

### B. Immunisation phase Groups g1,g2,g3: clusters

The immunisation phase network shows a similar structure. However, the names of countries and provinces that group together differ almost entirely from those seen in the outbreak phase. The names corresponding to the identified clusters in the groups g1,g2, and g3 are listed in Tables S4, S5, and S4, respectively.

## II. FIGURES

### A. Networks at zero threshold and network’s overlap at *θ* = 0.5

We show the correlation networks at the zero thresholds, both for the outbreak and immunisation phase studied in this work, see Fig. S1. These networks contain all 255 nodes in the connected component and more links, specifically 15128 for the outbreak and 16850 for the immunisation network. They have a large density (*ρ* =0.467, and 0.52) and a high clustering coefficient (*< Cc >*=0.78, and 0.76) for the outbreak and immunisation period, respectively. Notably, the corresponding networks at zero thresholds already show a community structure with three communities, particularly prominent in the outbreak phase, which persists at the higher threshold values. Additionally, Fig. S2 illustrates the differences between the outbreak and immunisation network measured by the preserved links, as mentioned in the main text.

### B. Time series cycles, Hurst exponents, and multifractality

Figure S3 complements the main text Fig. 7, showing that the infection cycles occur in both phases; moreover, they are more pronounced in the immunisation phase, even though the selected countries have a respectively high percentage of the vaccinated population. The fluctuations around these cycles, shown in Fig. S5, have certain multifractal properties, especially for small-scale fluctuations (for negative *q* values), and they are specific for each considered country.

**Supplementary Table S1:** Outbreak phase: Clusters of countries/regions belonging to the community G1.

| G1c1 | G1c2 | g1c3 | G1c4 |
| --- | --- | --- | --- |
| Australia-Western-Australia<br>Canada-Nova-Scotia<br>China-Anhui; China-Beijing<br>China-Chongqing<br>China-Guangdong<br>China-Heilongjiang<br>China-Henan; China-Hunan<br>China-Jiangsu; China-Jiangxi<br>China-Shandong<br>China-Shanghai<br>China-Zhejiang<br>New Zealand<br>Niger; San-Marino; Tanzania<br>UK-Channel-Island | China-Hubei | Tajikistan | Australia-Capital-Territory<br>Australia-Northern-Territory<br>Australia-Tasmania; Barbados; Brunei<br>Canada-Newfoundland-a.Labrador<br>Canada-Prince-Edward-Island<br>China-Fujian; China-Gansu; China-Guangxi; China-Hainan<br>China-Hebei;China-Inner-Mongolia; China-Jilin<br>China-Liaoning; China-Ningxia; China-Shaanxi<br>China-Shanxi; China-Sichuan; China-Tianjin; China-Yunnan<br>Denmark-Faroe-Islands; Denmark-Greenland; Fiji<br>France-New-Caledonia; Grenada; Laos; Mauritius<br>Saint-Kitts-a.Nevis; Timor-Leste; UK-Anguilla<br>UK-Bermuda; UK-Cayman-Islands; UK-Falkland-Islands<br>UK-Isle-of-Man; UK-Montserrat |

**Supplementary Table S2:** Outbreak phase: Clusters of countries/regions belonging to the community *G*2.

| G2c1 | G2c2 | G2c3 | G2c4 | G2c5 |
| --- | --- | --- | --- | --- |
| Bangladesh<br>Chile<br>Germany<br>Italy<br>Kazakhstan<br>Pakistan<br>Saudi-Arabia<br>United-Kingdom | Brasil | Afghanistan; Algeria; Australia-N.South-Wales<br>Australia-Queensland; Australia-South-Australia<br>Azerbaijan; Bahrain; Belarus; Belgium; Benin; Bolivia<br>Cameroon; Canada-Ontario; Canada-Quebec<br>Central-African-Republic; Chad; China-Macau<br>Congo(Brazzaville); Congo(Kinshasa); Cote-d'Ivoire<br>Djibouti; Dominican-Republic; Egypt; El-Salvador<br>Eswatini; Finland; France-French-Guiana; Gabon; Gambia<br>Ghana; Guatemala; Haiti; Honduras; Kuwait; Kyrgyzstan<br>Liberia; Madagascar; Malawi; Malaysia; Mali; Mauritania<br>Nigeria; Oman; Panama; Papua-New-Guinea; Qatar<br>Rwanda; Senegal; Seychelles; Sierra-Leone; Singapore<br>Somalia; South-Sudan; Sri-Lanka; Sudan; Suriname<br>Sweden; Taiwan*; Thailand; United-Arab-Emirates<br>UK-Turks-a.Caicos-Islands;Uzbekistan; Yemen; Zambia | Russia<br>South-Africa | Peru |

**Supplementary Table S3:**
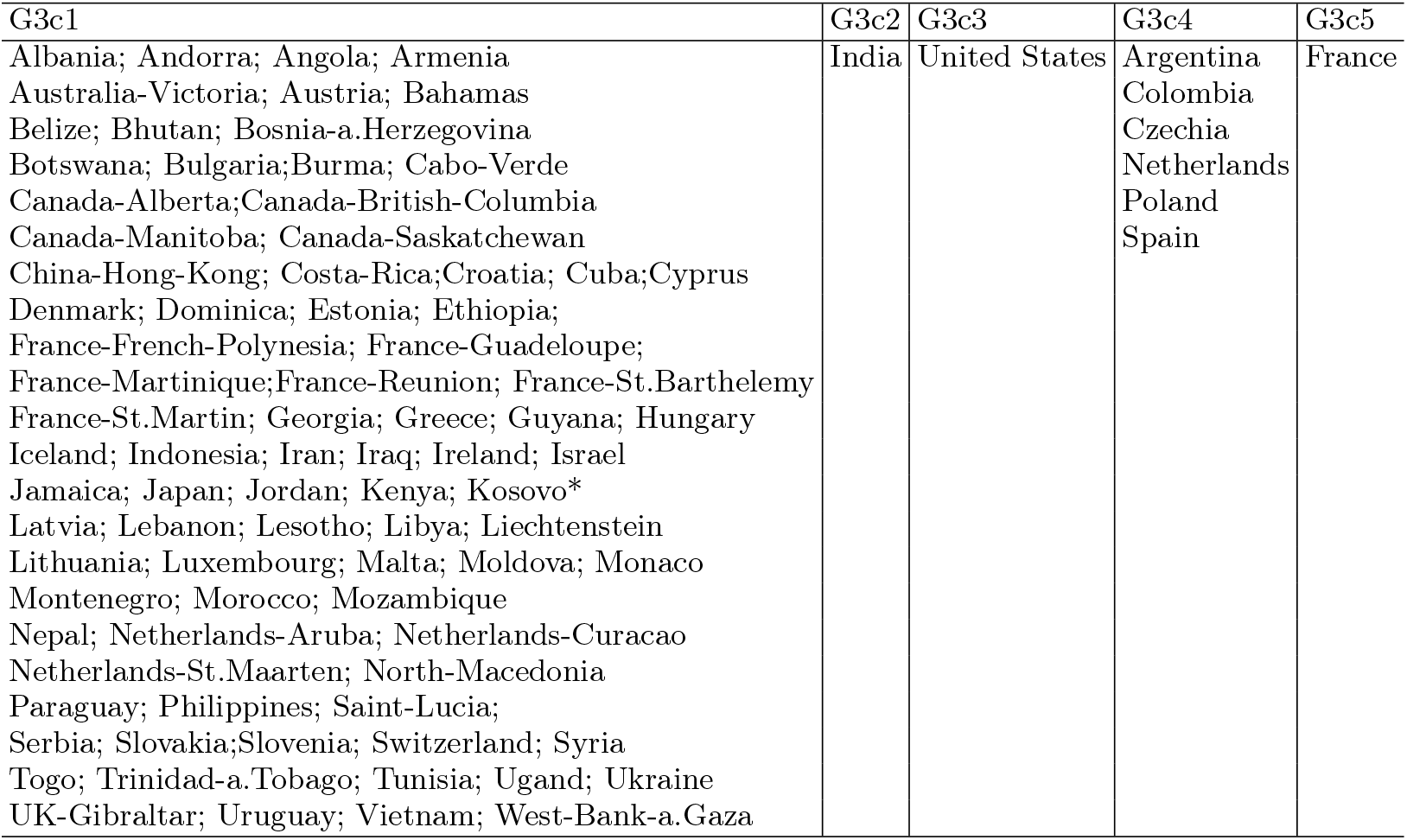
Outbreak phase: Clusters of countries/regions belonging to the community *G*3.

| G3c1 | G3c2 | G3c3 | G3c4 | G3c5 |
| --- | --- | --- | --- | --- |
| Albania; Andorra; Angola; Armenia<br>Australia-Victoria; Austria; Bahamas<br>Belize; Bhutan; Bosnia-a.Herzegovina<br>Botswana; Bulgaria;Burma; Cabo-Verde<br>Canada-Alberta;Canada-British-Columbia<br>Canada-Manitoba; Canada-Saskatchewan<br>China-Hong-Kong; Costa-Rica;Croatia; Cuba;Cyprus<br>Denmark; Dominica; Estonia; Ethiopia;<br>France-French-Polynesia; France-Guadeloupe;<br>France-Martinique;France-Reunion; France-St.Barthelemy<br>France-St.Martin; Georgia; Greece; Guyana; Hungary<br>Iceland; Indonesia; Iran; Iraq; Ireland; Israel<br>Jamaica; Japan; Jordan; Kenya; Kosovo*<br>Latvia; Lebanon; Lesotho; Libya; Liechtenstein<br>Lithuania; Luxembourg; Malta; Moldova; Monaco<br>Montenegro; Morocco; Mozambique<br>Nepal; Netherlands-Aruba; Netherlands-Curacao<br>Netherlands-St.Maarten; North-Macedonia<br>Paraguay; Philippines; Saint-Lucia;<br>Serbia; Slovakia;Slovenia; Switzerland; Syria<br>Togo; Trinidad-a.Tobago; Tunisia; Ugand; Ukraine<br>UK-Gibraltar; Uruguay; Vietnam; West-Bank-a.Gaza | India | United States | Argentina<br>Colombia<br>Czechia<br>Netherlands<br>Poland<br>Spain | France |

**Supplementary Table S4:** Immunisation phase: Clusters of countries/regions belonging to the community *g*1.

| g1c1 | g1c2 | g1c3 | g1c4 |
| --- | --- | --- | --- |
| Afghanistan; Bahrain; Bhutan; Bolivia; Cabo-Verde; Cambodia<br>Canada-Manitoba; Canada-Nova-Scotia; Canada-Ontario; Canada-Yukon<br>Chile; China-Anhui; China-Guangdong; China-Sichuan<br>Congo(Kinshasa); Dominican-Republic; Ecuador; Egypt; Eritrea;<br>Haiti; Kuwait; Kyrgyzstan; Liberia; Madagascar; Maldives<br>Namibia; Nepal; Oman; Panama; Paraguay; Saudi-Arabia;<br>Seychelles; Sierra-Leone; Taiwan*; Trinidad-a.Tobago;<br>Tunisia; United-Arab-Emirates; Uruguay; Zambia | India | Brasil | Argentina<br>Colombia<br>South-Africa |

**Supplementary Table S5:** Immunisation phase: Clusters of countries/regions belonging to the community *g*2.

| g2c1 | g2c2 | g2c3 | g2c4 | g2c5 |
| --- | --- | --- | --- | --- |
| Albania; Andorra; Armenia; Australia-Northern-Territory<br>Austria; Belarus; Belgium; Bosnia-a.Herzegovina<br>Bulgaria; Burkina-Faso; Cameroon; Canada-Alberta<br>Canada-British-Columbia; Canada-Newfoundland-a.Labrado<br>Canada-Quebec; Canada-Saskatchewan<br>Chad; China-Hong-Kong; China-Jiangxi; Comoros<br>Cote-d'Ivoire; Croatia; Djibouti; Estonia; Ethiopia<br>France-Mayotte; Gabon; Jamaica; Latvia; Lithuania<br>Luxembourg; Mali; Malta; Moldova; Monaco<br>Montenegro; Netherlands-Curacao; North-Macedonia<br>Norway; Pakistan; Qatar; Romania<br>San-Marino; Sao-Tome-a.Principe; Slovakia; Slovenia;<br>Somalia; South-Sudan; Sudan; Switzerland<br>Syria; UK-Falkland-Islands(Malvinas)<br>UK-Turks-a.Caicos-Islands; West-Bank-a.Gaza; Yemen | France | Czechia<br>Hungary<br>Jordan<br>Netherlands<br>Peru<br>Serbia<br>Sweden<br>Ukraine | Turkey | Germany<br>Italy<br>Poland |

**Supplementary Table S6:** Immunisation phase: Clusters of countries/regions belonging to the community *g*3.

| g3c1 | g3c2 | g3c3 | g3c4 | g3c5 |
| --- | --- | --- | --- | --- |
| Algeria; Angola; Australia-Capital-Territory<br>Australia-New-South-Wales; Australia-Queensland<br>Australia-Tasmania; Australia-Victoria<br>Azerbaijan; Bahamas; Belize; Benin<br>Botswana; Brunei; Burma; Burundi<br>Canada-Northwest-Territories<br>China-Fujian; China-Henan; China-Hubei<br>China-Hunan; China-Jiangsu; China-Shandong<br>China-Shanghai; China-Tianjin; China-Yunnan<br>Costa-Rica; Cuba; Cyprus<br>Denmark-Faroe-Islands; Denmark-Greenland<br>Dominica; Eswatini; Fiji<br>France-French-Guiana; France-French-Polynesia<br>France-Guadeloupe; France-Martinique<br>France-New-Caledonia; France-Reunion<br>Gambia; Georgia; Grenada; Guatemala<br>Guinea-Bissau; Guyana; Honduras; Iceland<br>Iraq; Ireland; Israel; Kazakhstan<br>Korea-South; Laos; Lesotho; Libya<br>Mauritania; Mauritius; Mongolia; Morocco<br>Netherlands-Aruba; Netherlands-St.Maarten<br>New-Zealand; Nicaragua; Portugal<br>Sain-Kitts-a.Nevis; Saint-Lucia; Singapore<br>Sri-Lanka; Suriname; Tajikistan; Timor-Leste<br>UK-Anguilla; UK-Cayman-Islands; UK-Gibraltar<br>UK-Isle-of-Man; Uzbekistan | United-States | Iran<br>United-Kingdom | Bangladesh<br>Japan<br>Malaysia<br>Mexico<br>Philippines<br>Russia<br>Thailand<br>Vientnam | Spain |

**Supplementary Figure S1:**
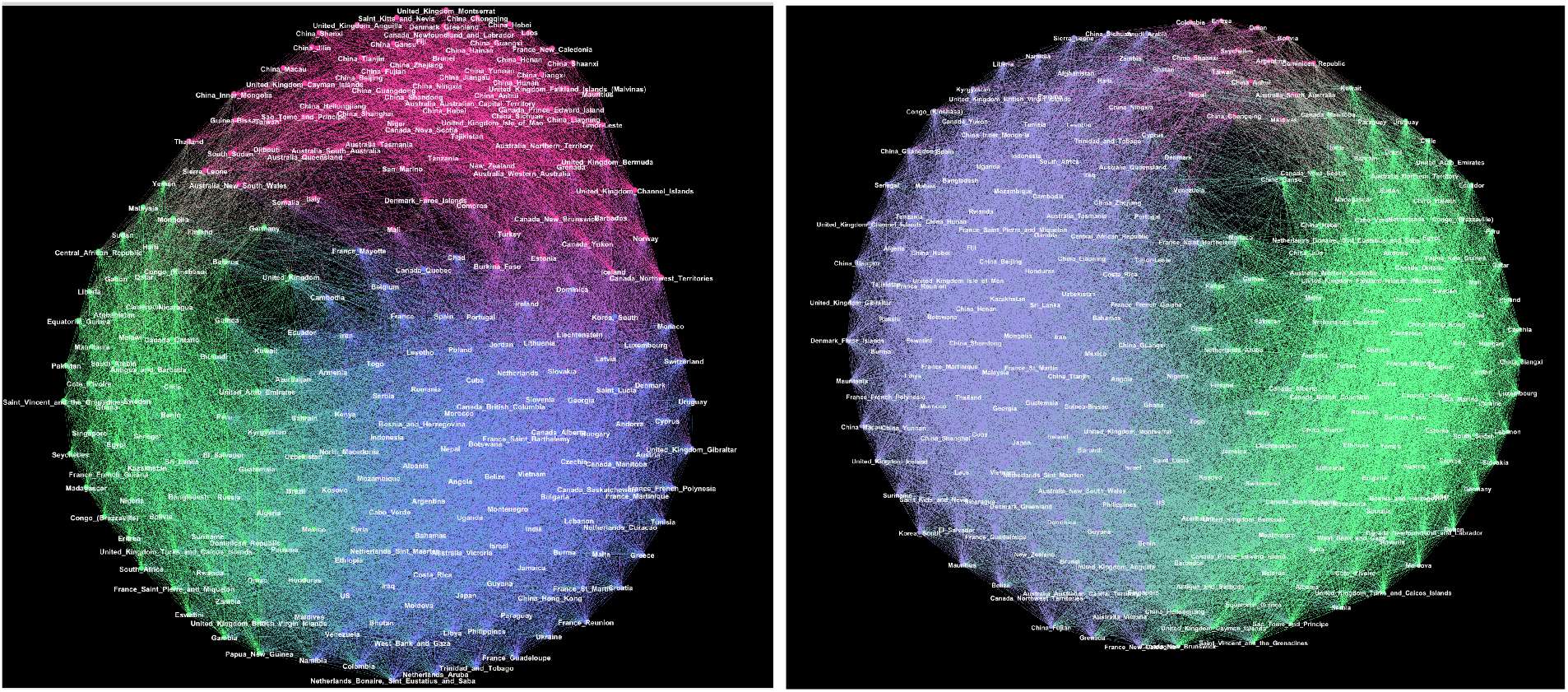
The networks containing all positive correlations (threshold *θ* = 0) for the outbreak (left) and immunisation phase (right); communities are indicated by a different colour.

**Supplementary Figure S2:**
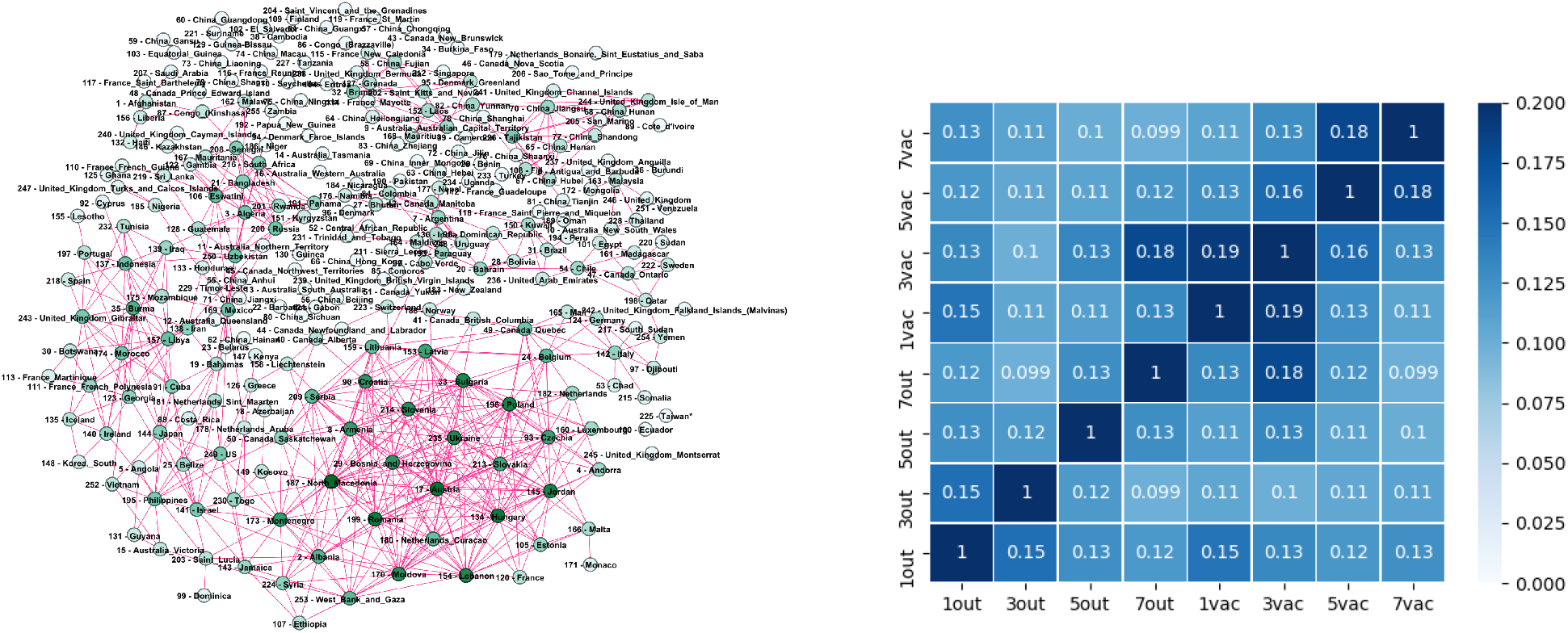
Left: The edges appearing in both the outbreak and immunisation periods networks with the threshold *θ* = 0.5, depicted in Fig. 3 in the main text. Right: The overlap level measured by the Jaccard index between networks mapping the successive two-months periods during the outbreak and immunisation phase.

**Supplementary Figure S3:**
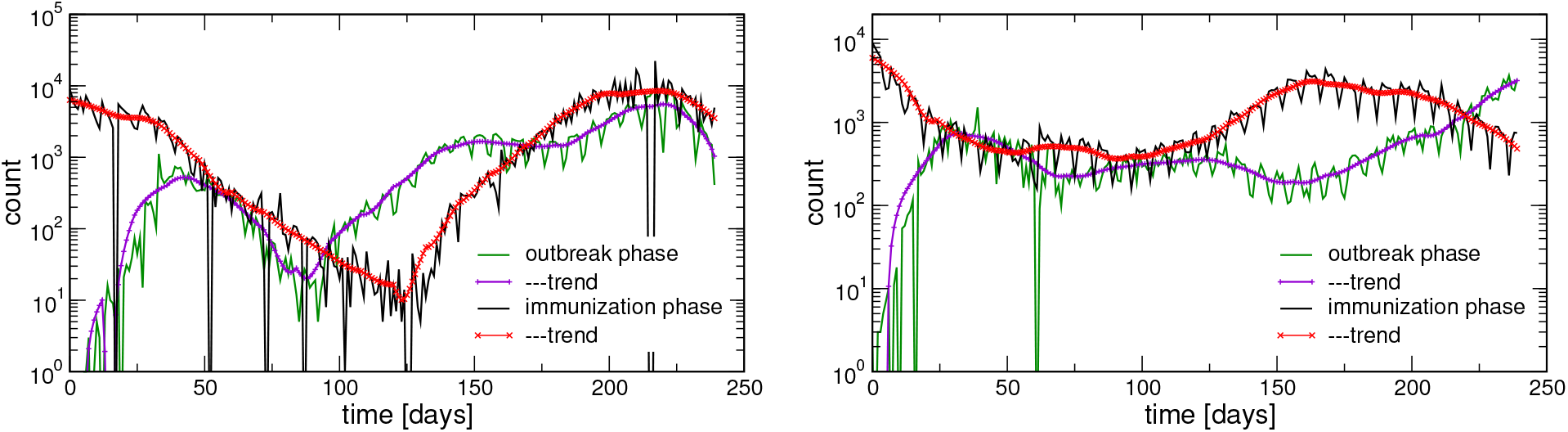
Comparisons of the outbreak and immunisation phases. Cyclic trends in the incidence rates data for two countries with a significant immunisation coverage (Israel, left, and Portugal, right).

**Supplementary Figure S4:**
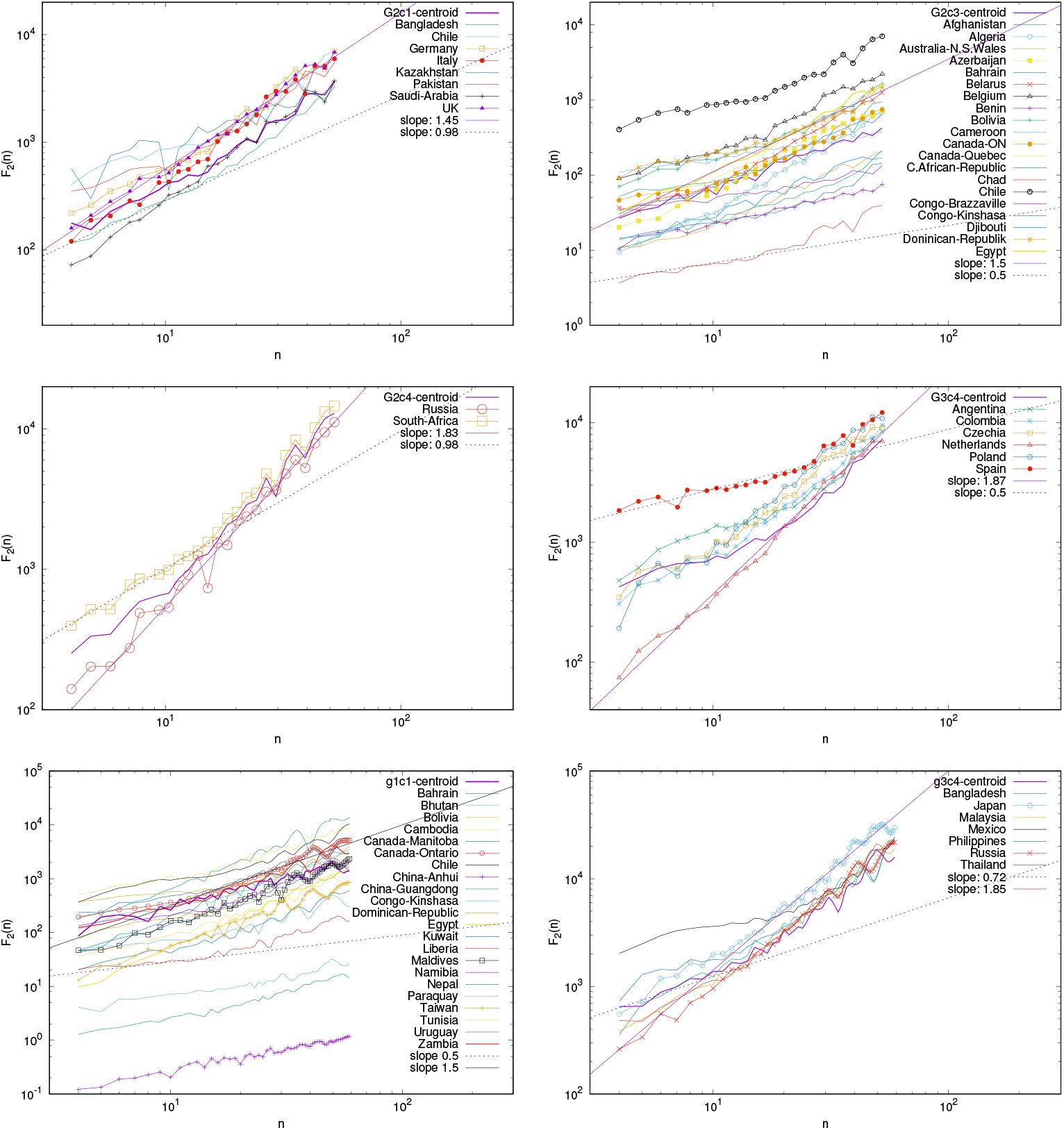
Fluctuation function *F*_2_(*n*) vs *n* for several identified clusters G2c1, G2c3 (part), G2c4, and G3c4 clusters in the outbreak phase, and g1c1 (part) and g3c4 clusters in the immunisation phase, mentioned in the main text.

**Supplementary Figure S5:**
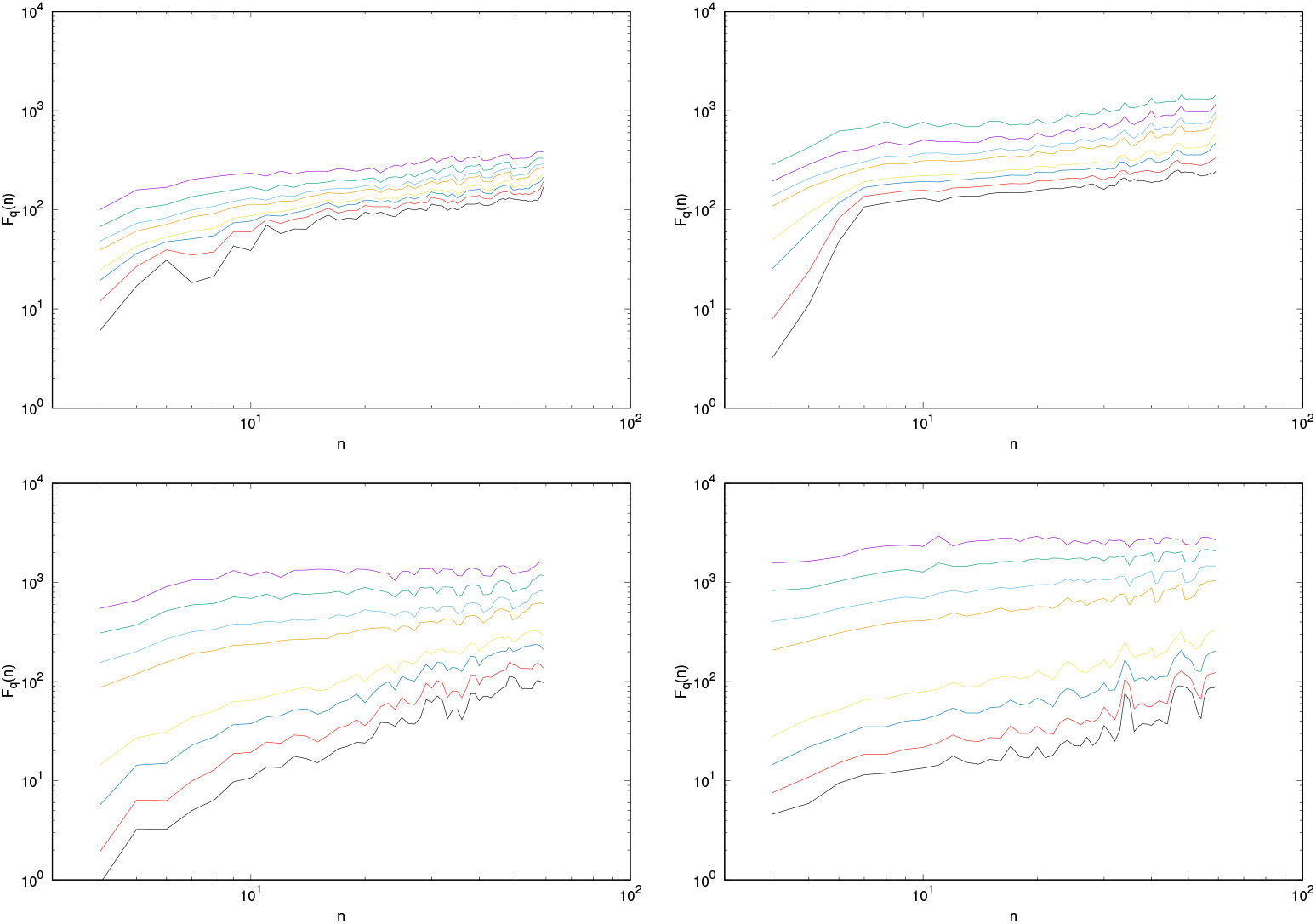
Fluctuations around the cyclic trend differ in the outbreak and immunisation phase. Two examples of the multifractal function *F*_*q*_ (*n*) vs interval length *n* for the time series data in the outbreak phase (left) and vaccination (right): top panels are for Portugal, bottom panels for Israel time-series data. In each panel, top to bottom lines are for *q* =4,2,1 0.5, -0.5,-1,-2, -4, respectively.

